# Targeted Pulsed Radio Frequency (PRF) Stimulation in the Management of Diabetic Peripheral Neuropathy: A Randomized, Single-Blind, Placebo-Controlled Trial

**DOI:** 10.64898/2026.08.07.26359945

**Authors:** Lukas D. Linde, Phyllis P. Berger, Stanley S. Landau, Elena Libhaber, Paul Potgieter, Petra van Blerk, Corlius F. Birkill

## Abstract

**Objective:** To evaluate the clinical efficacy of non-invasive electrical pulsed radiofrequency (PRF) stimulation on diagnostic thresholds and subjective pain in chronic, pedal diabetic peripheral neuropathy (DPN).

**Methods:** A randomized, single-blind, placebo-controlled trial (ClinicalTrials.gov: NCT07725419) enrolled 92 patients with pedal DPN naive to PRF and scoring ≥ 4/10 on the Douleur Neuropathique 4 (DN4) test. Participants received either active PRF stimulation (n = 46) or a non-stimulating placebo (n = 46) applied bilaterally to the sciatic nerve in the popliteal fossa for 10 minutes per limb, once weekly for three weeks. The primary outcome was clinical neuropathic resolution (DN4 < 4). Secondary outcomes included subjective pain tracking via the Brief Pain Inventory-Short Form (BPI-SF) Worst Pain scale over a 6-month follow-up window.

Missing data were handled via Non-Responder Imputation (NRI). Longitudinal continuous trajectories were modeled using Linear Mixed-Effects Models (LMMs) adjusted for age, gender, and baseline medication use.

**Results:** In the Intention-to-Treat population (N = 92), a significant diagnostic responder effect occurred at 3 months, with 39.1% of active patients dropping below the diagnostic threshold for neuropathy (DN4 < 4) versus 19.6% of placebo controls (p = 0.039). For subjective pain, 47.7% of active patients achieved a Minimally Clinically Important Difference (≥ 3-point reduction) in BPI Worst Pain at 1 month compared to 19.4% of placebo controls (p = 0.008). Multivariable logistic regression identified active treatment as a significant independent predictor of clinical response (Adjusted OR = 4.86; 95% CI: 1.56 to 17.53; p = 0.010). Continuous LMM tracking confirmed a statistically significant treatment-by-timepoint interaction for BPI Worst Pain at 1 month (p = 0.046).

**Conclusion:** A brief, three-week course of non-invasive PRF stimulation serves as a safe, effective, non-pharmacological adjunct that aids in managing the diagnostic presentation of neuropathic pain and mitigates worst pain experiences in patients suffering from pedal DPN.

## Introduction

Diabetic peripheral neuropathy (DPN) is a chronic, symmetrical, length-dependent sensorimotor polyneuropathy that affects up to 50% of the diabetic population over their lifetime (Kurz et al., 2026; Hicks & Selvin, 2019). DPN is characterized by distal, symmetrical symptoms that frequently worsen at night, including numbness, tingling, burning, and sharp, electric-shock-like pain (Feldman et al., 2019). Due to its persistent and debilitating nature, painful DPN severely diminishes health-related quality of life, disrupts sleep, and significantly increases the risk of foot ulceration and amputation (Hicks & Selvin, 2019; Armstrong et al., 2017).

Furthermore, despite its widely recognized prevalence and heavy clinical burden, DPN remains notoriously inadequately treated across various clinical settings (Ziegler et al., 2022).

The initial approach to treating painful DPN tends to be pharmacological, relying on interventions such as tricyclic antidepressants, gabapentinoids, and serotonin-norepinephrine reuptake inhibitors (Ziegler et al., 2022; Sloan et al., 2021). However, these symptomatic therapies are often unsatisfactory, as they frequently provide inadequate pain relief and their use is heavily complicated by adverse side effects, such as dizziness, somnolence, and cognitive impairment (Feldman et al., 2019; Finnerup et al., 2010; Sloan et al., 2021). This leaves a substantial unmet clinical need, forcing many patients to discontinue treatment and seek alternative measures. Consequently, non-invasive neuromodulation modalities have been increasingly explored as an adjunct pain management solution. However, a recent comprehensive meta-analysis of these interventions demonstrated that while specific peripheral electrical techniques (such as TENS) can reduce neuropathic pain, peripheral electromagnetic interventions fail to yield significant analgesic effects (Zeng et al., 2020). Furthermore, the beneficial effects of these conventional non-invasive modalities are highly protocol-dependent, frequently requiring intensive, continuous clinical application to achieve and maintain meaningful relief (Zeng et al., 2020).

To address the limitations of conventional and strictly pharmacological treatments, PRF has emerged as a highly promising neuromodulatory intervention for neuropathic pain. Unlike continuous radiofrequency, PRF modulates nerve signaling and neuroimmune pathways without causing thermal tissue destruction (Sluijter et al., 2023). Mechanistically, emerging evidence suggests these non-destructive analgesic and regenerative effects are driven by targeted modulations of neuronal bioenergetics, specifically by acutely depolarizing the mitochondrial membrane to mitigate excitotoxic calcium influx and promoting sub-acute mitochondrial hyperplasia (Linde et al., 2026). Recent clinical evidence has validated the efficacy of PRF in managing treatment-resistant DPN, demonstrating significant improvements in pain severity, sleep duration, and analgesic reliance (Wang et al., 2024). Building on these principles, non-invasive PRF delivered via transcutaneous electrodes has recently been developed to eliminate the risks associated with percutaneous needle placement. Emerging randomized controlled trial data indicates that non-invasive PRF yields robust, statistically significant reductions in DPN pain scores compared to sham stimulation (Genç Perdecioğlu et al., 2025). However, while these initial non-invasive findings are highly encouraging, comprehensive longitudinal evaluations utilizing diagnostic assessments and multidimensional pain inventories over extended follow-up windows remain scarce.

To establish the clinical viability of this intervention, a randomized, single-blind, placebo-controlled trial was conducted to evaluate the efficacy of a non-invasive PRF device in patients with treatment-resistant pedal DPN. Treatment efficacy was evaluated based on a clinically meaningful reduction in the Douleur Neuropathique 4 (DN4) assessment as the primary outcome measure. Secondary endpoints tracked subjective pain severity and its functional impact utilizing the Brief Pain Inventory-Short Form (BPI-SF), focusing on reductions in worst pain intensity. It was hypothesized that a three-week course of active PRF stimulation would achieve superior diagnostic resolution of neuropathic pain via the DN4 assessment and deliver a sustained reduction in worst pain severity compared to a non-stimulating placebo.

## Methods

### Study Design and Ethical Approval

This study represents a comprehensive reanalysis of clinical trial data originally reported by Berger and Landau (Berger & Landau, 2020). While the original publication detailed the foundational clinical findings, the current reanalysis applies updated, robust longitudinal methodologies to further evaluate the treatment trajectory. A randomized, single-blind, placebo-controlled trial was conducted to evaluate the efficacy of a non-invasive PRF device on neuropathic pain symptoms in patients with pedal DPN. The study design was approved by the Human Research Ethics Committee (Medical) of the University of the Witwatersrand (Clearance number: M161037) prior to participant enrollment, and permission was granted by physicians at the Centre for Diabetes and Endocrinology (CDE) in Johannesburg and Pretoria. Following prospective ethical clearance, the trial was retrospectively registered with ClinicalTrials.gov (Identifier: NCT07725419). All patients provided written informed consent prior to participation.

### Participants and Randomization

Patients were screened telephonically and subsequently assessed for eligibility.

Inclusion criteria required a diagnosis of pedal DPN, a score of ≥ 4/10 on the DN4 assessment, and being naive to electrical PRF stimulation. Exclusion criteria included pregnancy, pacemakers, peripheral or central nerve stimulators, and metal implants in the knee or hip. A total of 92 patients were enrolled and randomized into either the Active treatment group (Group A, n=46) or the Placebo group (Group B, n=46) to accommodate anticipated attrition (Figure 1). The study cohort represented diverse socioeconomic backgrounds; participant-reported racial and ethnic demographics included Black, White, Indian, and Asian populations. All participant recruitment, interventions, and follow-up sessions took place from December 2016 until August 2019.

**Figure 1:**
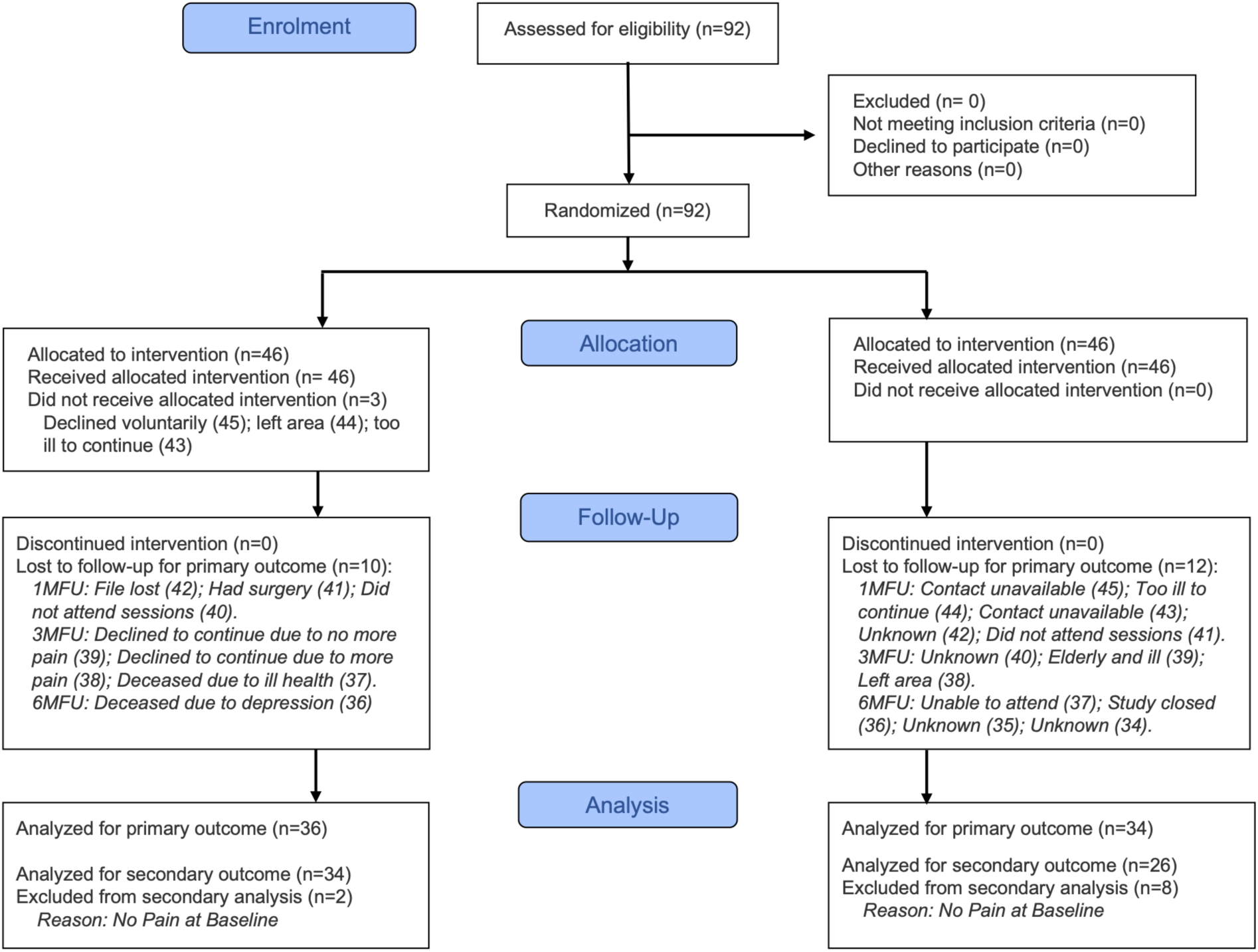
CONSORT Participant Flow Diagram. Flow of participants through randomized, single-blind, placebo-controlled trial evaluating non-invasive pulsed radio frequency (PRF) intervention for pedal diabetic peripheral neuropathy. Diagram tracks the progression of 92 randomized patients across the active treatment (n = 46) and placebo control (n = 46) groups from baseline allocation to the final six-month follow-up timepoint. Sample sizes for the primary analysis consist of the Intention-to-Treat (ITT, n = 92) and Per-Protocol (PP, n = 70) cohorts, while secondary pain analyses utilize the modified Intention-to-Treat (mITT, n = 80) and modified Per-Protocol (mPP, n = 60) cohorts, which exclude patients with a baseline BPI-Worst Pain score of 0.

### Blinding and Intervention

Due to the technical nature of the active PRF stimulation, which can induce visible muscle fasciculations to indicate correct nerve targeting, double blinding was not attainable. A single-blind design was employed. Participant blinding was rigorously maintained; patients were informed that due to the high-frequency nature of the stimulation and their varying neuropathic manifestations, they might or might not perceive the stimulation. Because all participants were naive to the intervention, this effectively prevented them from distinguishing between active and placebo assignments based on sensation alone. Four unblinded investigators administered the treatments across multiple centers. To mitigate performance and detection bias, the study relied strictly on standardized, patient-reported outcome measures.

During treatment, a nerve mapping device was used to locate the sciatic nerve in the popliteal fossa posteriorly at the knee, and the position was marked. The intervention was administered using a transcutaneous electrical pulsed radiofrequency device (NMS 460; Xavant Technology Pty Ltd). The device (Active or Placebo) was applied bilaterally for 10 minutes per limb, for a total treatment time of 20 minutes. The stimulation intensity was adjusted to elicit fasciculation (although visible response varied based on the severity of patient’s neuropathy) and recorded at each session to ensure patient comfort. The intervention was administered once weekly for three consecutive weeks, for a total of three treatment sessions.

### Outcome Measures

Assessments were conducted at baseline, immediately following the final (third) treatment, and at 1-, 3-, and 6-month follow-ups. The primary outcome, neuropathic diagnostic resolution, was assessed using the DN4 questionnaire, which consists of seven subjective symptom questions and three objective clinical tests (Bouhassira et al, 2005). Secondary outcomes focused on subjective pain severity and functional interference, which were measured using the Brief Pain Inventory-Short Form (BPI-SF). The BPI-SF captures worst, least, average, and present pain on an 11-point numerical rating scale ranging from 0 to 10, alongside a 7-item composite evaluating general activity, mood, walking ability, normal work, relations with others, sleep, and enjoyment of life (Cleeland & Ryan, 1994; Zelman et al., 2005). Additionally, longitudinal concurrent systemic analgesic medication use was tracked throughout the trial.

### Statistical Analysis

All statistical analyses were performed using R (R Core Team, 2024). To further ensure objectivity and eliminate investigator bias, data management and statistical analyses of the raw participant-level dataset were completed by an independent analyst completely removed from the original clinical data collection. To provide a conservative evaluation of efficacy, missing data were handled using Non-Responder Imputation (NRI), wherein any missing post-treatment value was automatically classified as a non-response.

The analysis was executed across predefined cohorts based on the updated CONSORT flow diagram (Hopewell et al., 2025): the Intention-to-Treat (ITT, N=92) and Per-Protocol (PP, N=70) populations for the primary diagnostic endpoint (DN4), and modified Intention-to-Treat (mITT, N=80) and modified Per-Protocol (mPP, N=60) populations for the secondary subjective pain endpoints (BPI). The modified cohorts specifically excluded patients with a baseline BPI-Worst Pain score of 0 to prevent artificial floor effects.

Responder analyses were evaluated using unadjusted Pearson’s Chi-square tests (with corresponding 95% Confidence Intervals for risk differences). Positive clinical responses were defined independently for each metric using established thresholds. For the primary outcome, a responder was defined as dropping below the diagnostic threshold for neuropathy on the DN4 (< 4) (Spallone et al., 2012). For the secondary outcome, a responder was defined as achieving a Minimally Clinically Important Difference (MCID) of a ≥ 3-point reduction from baseline on the BPI-Worst Pain scale (Marcus et al., 2018). Because subjective pain severity is highly susceptible to demographic and pharmacological confounding, the BPI-Worst Pain responder analysis was further evaluated using multivariable logistic regression, adjusting for age, gender, baseline pain severity, and baseline medication use, to confirm the independent efficacy of the intervention. Categorical changes in longitudinal medication use from baseline to the 6-month follow-up were evaluated using McNemar’s test for paired proportions.

For continuous longitudinal tracking, the BPI metrics (Worst, Least, Average, Present, and the Interference Composite) were analyzed using Linear Mixed-Effects Models (LMMs). To account for potential confounding over the 6-month trajectory, the models were adjusted for age, gender, and baseline medication use, with Treatment Group and Timepoint fit as interacting fixed effects, and participants as a random intercept. The sample size for this cohort was originally calculated *a priori* to detect a clinically meaningful difference in the primary diagnostic endpoint (DN4). Because the trial was not explicitly powered for complex multivariable longitudinal analyses of secondary endpoints, the adjusted LMMs were deliberately restricted to a minimal set of clinically critical covariates (age, gender, and baseline medication use) to prevent model overfitting and preserve statistical power. Significance was established at a two-sided alpha level of p < 0.05 for all statistical tests.

## Results

### Baseline Characteristics

A total of 92 patients with pedal diabetic peripheral neuropathy were assessed for eligibility, enrolled, and randomized into the trial, comprising the Intention-to-Treat (ITT) cohort (Active: n = 46; Placebo: n = 46) (Figure 1). The Per-Protocol (PP) cohort, which included patients who completed all required treatment and follow-up visits, consisted of 70 patients (Active: n = 36; Placebo: n = 34). Baseline demographics, clinical characteristics, baseline pain severity, and concurrent systemic medication use were summarized for both groups (Table 1).

**Table 1:** Baseline Demographics and Clinical Characteristics.

| Characteristic | Active (n = 46) | Placebo (n = 46) |
| --- | --- | --- |
| Age (Mean ± SD) | 63.8 ± 9.4 | 59.1 ± 9.6 |
| Sex, n (%) |  |  |
| Female | 17 (37.0%) | 15 (32.6%) |
| Male | 29 (63.0%) | 31 (67.4%) |
| Baseline DN4 (Mean ± SD) | 6.4 ± 1.6 | 6.2 ± 1.6 |
| Baseline BPI Worst Pain (Mean ± SD) | 6.9 ± 2.3 | 5.6 ± 3.5 |
| Baseline Medication Use, n (%) |  |  |
| Yes | 29 (63.0%) | 24 (52.2%) |
| No | 17 (37.0%) | 22 (47.8%) |

### Primary Outcome: Neuropathic Diagnostic Resolution (DN4)

For the responder analysis of the DN4 primary outcome measure, a positive responder was classified as dropping below the diagnostic threshold for neuropathy (DN4 < 4). There was a significant responder effect for the DN4 assessment 3 months after intervention in the ITT cohort (n = 92), with 39.1% of responders in the active treatment group compared to 19.6% of responders in the placebo group (p = 0.039; Table 2). For the PP analysis, there was a significant responder effect for the DN4 immediately following treatment, with 66.7% of responders in the active treatment group compared to 32.4% of responders in the placebo group (p = 0.004; Table 2).

**Table 2:** Responder Analysis (DN4 score < 4).

| Timepoint | Intent-to-Treat Analysis (ITT) (N=92) |  |  |  | Per-Protocol Analysis (PP) (N=70) |  |  |  |
| --- | --- | --- | --- | --- | --- | --- | --- | --- |
|  | Active | Placebo | Risk Diff | P-Value | Active | Placebo | Risk Diff | P-Value |
| **Post-Tx** | 24/46<br>(52.2%) | 18/46<br>(39.1%) | 13.0%<br>[-7.1,<br>33.2] | 0.209 | 24/36<br>(66.7%) | 11/34<br>(32.4%) | 34.3%<br>[12.3,<br>56.3] | **0.004\*** |
| **1 Month** | 17/46<br>(37.0%) | 12/46<br>(26.1%) | 10.9%<br>[-8.0,<br>29.7] | 0.262 | 15/36<br>(41.7%) | 10/34<br>(29.4%) | 12.3%<br>[-10.0,<br>34.5] | 0.285 |
| **3 Months** | 18/46<br>(39.1%) | 9/46<br>(19.6%) | 19.6%<br>[1.4,<br>37.7] | **0.039\*** | 16/36<br>(44.4%) | 8/34<br>(23.5%) | 20.9%<br>[-0.7,<br>42.5] | 0.065 |
| **6 Months** | 16/46<br>(34.8%) | 9/46<br>(19.6%) | 15.2%<br>[-2.7,<br>33.1] | 0.101 | 15/36<br>(41.7%) | 9/34<br>(26.5%) | 15.2%<br>[-6.7,<br>37.1] | 0.181 |

Statistical Test: Pearson Chi-Square Test of Independence. In the ITT analysis, all participants with missing data (dropouts/exclusions) were imputed as non-responders.

### Secondary Outcome: Subjective Pain Severity (BPI)

For the responder analysis of the secondary outcome measure, BPI Worst Pain, a positive responder was classified as achieving a ≥ 3-point reduction from baseline. In the mITT cohort (n = 80), there was a significant responder effect at 1 month, with 47.7% of responders in the active group compared to 19.4% of responders in the placebo group (p = 0.008; Table 3). In the mPP cohort (n = 60), there was also a significant responder effect at 1 month, with 52.9% of responders in the active group compared to 26.9% of responders in the placebo group (p = 0.043; Table 3).

**Table 3:** Responder Analysis (BPI Worst Pain Severity Scores).

| Timepoint | Modified Intent-to-Treat Analysis (mITT) (N=80) |  |  |  | Modified Per-Protocol Analysis (mPP) (N=60) |  |  |  |
| --- | --- | --- | --- | --- | --- | --- | --- | --- |
|  | Active | Placebo | Risk Diff | P-Value | Active | Placebo | Risk Diff | P-Value |
| **Post-Tx** | 15/44<br>(34.1%) | 11/36<br>(30.6%) | 3.5%<br>[-17.0,<br>24.1] | 0.737 | 12/34<br>(35.3%) | 6/26<br>(23.1%) | 12.2%<br>[-10.6,<br>35.0] | 0.306 |
| **1 Month** | 21/44<br>(47.7%) | 7/36<br>(19.4%) | 28.3%<br>[8.7,<br>47.9] | **0.008\*** | 18/34<br>(52.9%) | 7/26<br>(26.9%) | 26.0%<br>[2.1,<br>49.9] | **0.043\*** |
| **3 Months** | 15/44<br>(34.1%) | 13/36<br>(36.1%) | -2.0%<br>[-23.1,<br>19.0] | 0.851 | 13/34<br>(38.2%) | 13/26<br>(50.0%) | -11.8%<br>[-37.0,<br>13.5] | 0.362 |
| **6 Months** | 14/44<br>(31.8%) | 10/36<br>(27.8%) | 4.0%<br>[-16.0,<br>24.1] | 0.695 | 13/34<br>(38.2%) | 10/26<br>(38.5%) | -0.2%<br>[-25.1,<br>24.6] | 0.986 |

Statistical Test: Pearson Chi-Square Test of Independence. In the mITT analysis, all participants with missing data (dropouts/exclusions) were imputed as non-responders.

A multivariable logistic regression of BPI Worst Pain adjusting for age, gender, baseline pain severity, and baseline medication use confirmed active treatment as a significant independent predictor of response to treatment at 1 month in the mITT cohort (Adjusted OR = 4.86; 95% CI: 1.56 to 17.53; p = 0.010; Supplementary Table 1). In the mPP cohort, the fully adjusted model did not yield a significant treatment effect at 1 month (Adjusted OR = 3.65; 95% CI: 0.98 to 16.14; p = 0.066; Supplementary Table 1).

### Continuous Longitudinal Tracking (LMM)

The continuous Linear Mixed-Effects Models (LMMs) over the 6-month trajectory corroborated the responder analysis for BPI outcomes. While both cohorts demonstrated an overall improvement in BPI Worst Pain over time, the active treatment group exhibited a statistically significant treatment-by-timepoint interaction exclusively at the 1-month follow-up. This interaction remained significant after adjusting for age, gender, and baseline medication use, indicating a steeper decline in BPI Worst Pain at 1 month follow-up compared to placebo (Adjusted Difference = -1.70; p = 0.046; Figure 2, Table 4). Conversely, treatment-by-timepoint interaction terms for BPI Average Pain, Least Pain, Present Pain, and the Interference Composite did not reach statistical significance at any timepoint (Table 4). Importantly, the unadjusted and fully adjusted models yielded comparable point estimates and standard errors (Supplementary Table 2). This indicates that the inclusion of the selected covariates did not overfit the model or substantially compromise statistical power, confirming that demographic factors and baseline medication use were not the primary drivers of the longitudinal pain trajectories.

**Figure 2:**
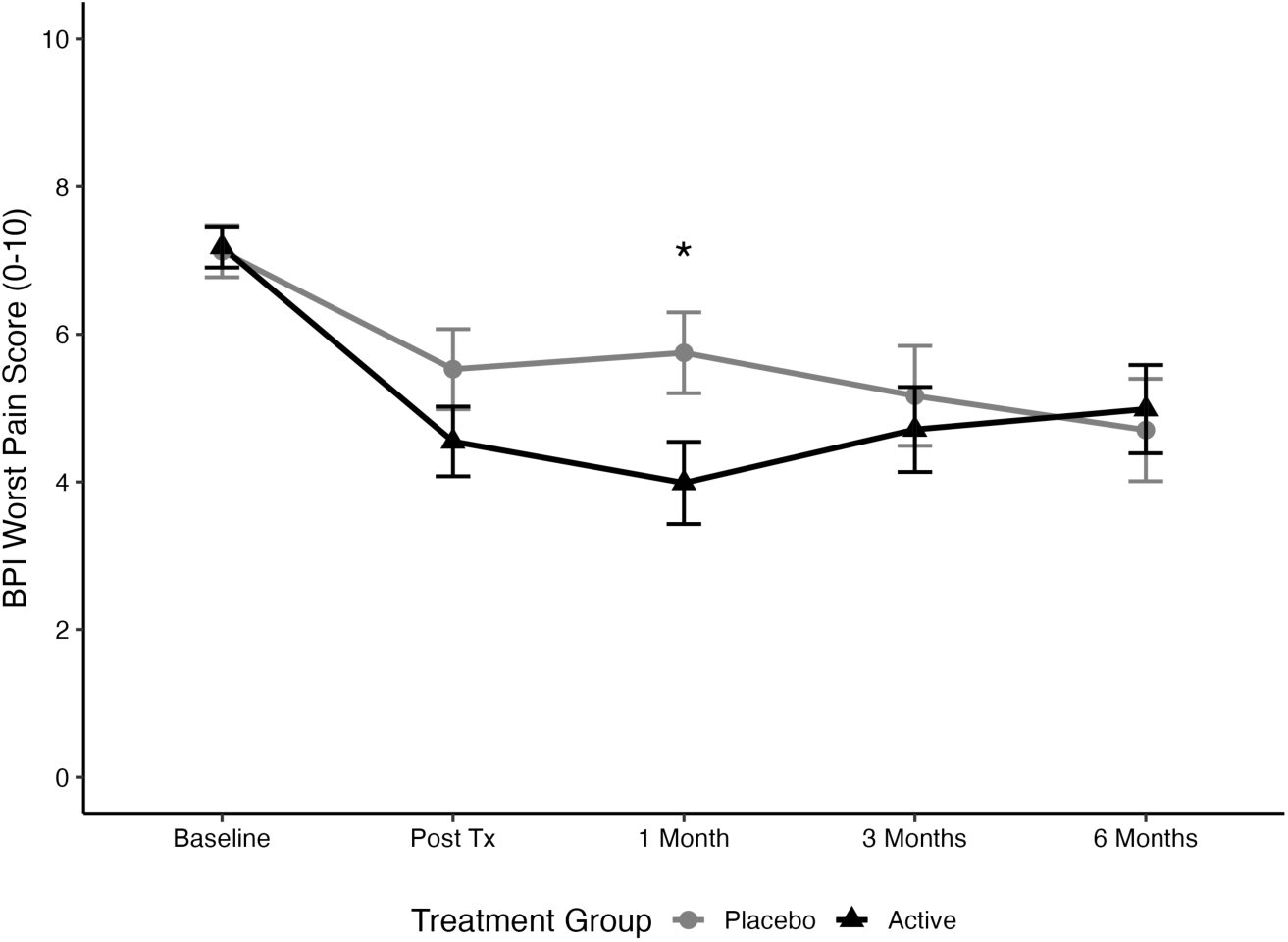
Longitudinal Trajectory of Brief Pain Inventory (BPI) Worst Pain Over 6 Months. Mean and standard errors for Brief Pain Inventory (BPI) Worst Pain tracked at baseline, immediately post-treatment (3 weeks), and at 1-, 3-, and 6-month follow-up timepoints for the modified Intention-to-Treat (mITT) cohort (n = 80; Active: n = 44, Placebo: n = 36). Linear Mixed-Effects Model (LMM) analysis adjusted for age, gender, and baseline medication use demonstrated a significant treatment-by-timepoint interaction at the 1-month follow-up index (p = 0.046), reflecting a greater reduction in worst pain severity for the active PRF treatment group compared to the placebo group. Corresponding responder rates for the mITT (n = 80) and modified Per-Protocol (mPP, n = 60; Active: n = 34, Placebo: n = 26) cohorts are detailed in text.

**Table 4:** Continuous Outcomes (Linear Mixed Model Interactions).

| Endpoint & Timepoint | Adjusted Difference (Active vs. Placebo)* | t-value | p-value |
| --- | --- | --- | --- |
| **BPI Worst Pain** |  |  |  |
| Post-Treatment | -0.96 | -1.17 | 0.242 |
| 1 Month | -1.70 | -2.01 | **0.046\*** |
| 3 Months | -0.52 | -0.61 | 0.541 |
| 6 Months | 0.04 | 0.04 | 0.964 |
| **BPI Least Pain** |  |  |  |
| Post-Treatment | 0.51 | 0.93 | 0.354 |
| 1 Month | 0.40 | 0.70 | 0.486 |
| 3 Months | 0.80 | 1.40 | 0.162 |
| 6 Months | 1.00 | 1.71 | 0.088 |
| **BPI Average Pain** |  |  |  |
| Post-Treatment | -0.64 | -0.93 | 0.353 |
| 1 Month | -1.30 | -1.83 | 0.068 |
| 3 Months | -0.22 | -0.30 | 0.762 |
| 6 Months | -0.02 | -0.03 | 0.979 |
| **BPI Present Pain** |  |  |  |
| Post-Treatment | -1.30 | -1.77 | 0.078 |
| 1 Month | -1.07 | -1.42 | 0.158 |
| 3 Months | -0.36 | -0.47 | 0.641 |
| 6 Months | -0.98 | -1.25 | 0.212 |
| **BPI Interference Composite** |  |  |  |
| Post-Treatment | -0.44 | -0.73 | 0.466 |
| 1 Month | -0.80 | -1.29 | 0.197 |
| 3 Months | -0.19 | -0.31 | 0.757 |
| 6 Months | -0.24 | -0.38 | 0.707 |

*A negative Adjusted Difference indicates a greater reduction in the pain score for the Active group compared to Placebo. All models adjusted for age, gender, and baseline medication use.*

### Safety and Concurrent Treatments

There were zero procedure- or device-related adverse events reported during the trial. Tracking of concurrent systemic analgesic use demonstrated a statistically significant decline in both cohorts over the 6-month follow-up period. Analgesic utilization in the active group decreased from 63.0% at baseline to 37.0% at 6 months (p = 0.014), and in the placebo control group decreased from 52.2% at baseline to 23.9% at 6 months (p = 0.002; Supplementary Table 3).

## Discussion

The primary objective of this randomized, single-blind, placebo-controlled trial was to determine whether a short course of non-invasive PRF stimulation, applied bilaterally to the sciatic nerve in the popliteal fossa, could produce a statistically significant and clinically meaningful reduction in pain and symptoms for patients suffering from chronic pedal DPN. The primary findings demonstrate a significant treatment effect, evidenced by a significant proportion of patients in the active treatment group achieving diagnostic resolution of their neuropathic pain presentation, successfully dropping below the established neuropathic threshold on the DN4 assessment. The clinical significance of this diagnostic resolution is highlighted by the mechanics of the DN4 tool itself, which integrates both subjective symptoms (burning, painful cold, and electric shocks) and objective physical signs (tingling, numbness, and touch hypoesthesia). This comprehensive evaluation is critical, as many DPN patients present with a severe combination of sensory deficits rather than isolated pain. Over time, as neuropathy progresses, patients often report a paradoxical decrease in acute pain that masks a profound worsening of sensory loss, a deceptive clinical state that frequently prevents individuals from recognizing localized foot injuries until catastrophic complications occur (Feldman et al., 2019). As such, treatments capable of modifying both the painful and non-painful somatosensory deficits of DPN are vital to improving long-term patient outcomes.

Importantly, the overall clinical efficacy demonstrated in this updated analysis corroborates the foundational findings of the original clinical trial (Berger & Landau, 2020). Both the original publication and the current reanalysis reach the same fundamental conclusion regarding the beneficial effect of PRF in pedal diabetic neuropathy. While this present evaluation applied stricter missing data handling through Non-Responder Imputation and utilized more conservative covariate adjustments for longitudinal modeling, the overarching clinical trajectory remains unchanged, confirming PRF as a reliable adjunct for mitigating neuropathic symptoms in those living with pedal diabetic peripheral neuropathy.

Evaluating subjective pain using the BPI highlighted a highly targeted clinical response isolated specifically to Worst Pain outcome, a likely surrogate for breakthrough hyperalgesia. While continuous longitudinal linear mixed-effects modelling (LMM) revealed that the interaction terms for Average Pain, Least Pain, and Present Pain did not achieve independent statistical significance over the full six-month trajectory, the active cohort experienced a significant, clinically meaningful mitigation Worst Pain experiences at the one-month follow-up. This selective impact on peak pain intensity represents a distinct therapeutic asset rather than a limitation. Because PRF stimulation does not induce a generalized, blunt anesthetic effect that numbs the extremities, it preserves remaining protective sensation in the vulnerable diabetic foot while successfully blunting the breakthrough hyperalgesia that heavily disrupts sleep and patient quality of life. Mechanistically, this aligns with recent literature suggesting that PRF stimulation mitigates acute excitotoxic calcium influx and modulates neuronal bioenergetics without inducing the thermal tissue destruction or indiscriminate neural blockade associated with continuous radiofrequency ablation (Sluijter et al., 2023; Linde et al., 2026).

These results address a critical gap in the current landscape of non-invasive neuromodulation. While conventional peripheral electromagnetic and electrical interventions, such as transcutaneous electrical nerve stimulation (TENS), can provide symptomatic relief, their analgesic effects are often temporary and heavily dependent on continuous, daily application (Zeng et al., 2020). In contrast, recent trials investigating both invasive and non-invasive PRF have demonstrated more robust and sustained neuromodulatory capabilities in treatment-resistant cohorts (Wang et al., 2024; Genç Perdecioğlu et al., 2025). The present study corroborates and extends these findings by demonstrating that even a highly abbreviated, three-week course of transcutaneous PRF stimulation can fundamentally alter the neuropathic diagnostic presentation and mitigate peak pain severity well beyond the active treatment window.

The reported findings also address the problematic role of concurrent pharmacological treatments, a ubiquitous confounding variable in non-pharmacological neuromodulation trials. Patients entered this study with long-standing DPN managed by a heterogenous array of systemic medications, analgesics, anticonvulsants, and antidepressants, frequently taken in variable combinations. Documenting these regimens presents clinical challenges due to intermittent patient compliance, poor memory regarding sporadic usage, and abrupt drug discontinuations driven by intolerable side effects. While standard unadjusted evaluations can easily become diluted by this extreme pharmacological heterogeneity, our fully adjusted LMMs and multivariable logistic regression isolated an independent treatment effect. The analysis demonstrated that active PRF stimulation achieved an adjusted odds ratio of nearly five for successful pain reduction at one month, an effect that persisted independent of baseline pharmacological covariates.

Several limitations must be considered when evaluating these clinical findings. The requirement to actively titrate the PRF stimulation intensity to monitor for motor fasciculations during sciatic nerve targeting fundamentally precluded a double-blind study design, leaving the treating investigators unblinded. To limit detection and performance bias, the trial relied strictly on standardized, patient-reported metrics where data were directly generated by masked participants, alongside an independent data management and analysis protocol. Furthermore, sample attrition naturally occurred across the six-month follow-up period, driven primarily by overlapping medical conditions and loss of contact with aging participants. Finally, while the study was adequately powered for its primary endpoints, the statistical margins of the secondary analyses must be interpreted contextually. The borderline significance in the Per-Protocol responder analysis (p = 0.043) and the wide confidence intervals in the Intention-to-Treat framework are largely a reflection of the highly conservative Non-Responder Imputation method used for missing data, combined with the inherent clinical heterogeneity of subjective pain reporting in chronic DPN.

While a brief, three-week intervention protocol established a modest but clinically meaningful therapeutic signal, chronic DPN is a progressive neurological pathology that likely demands a more continuous clinical dosing strategy. Prior clinical observations indicate that a brief three-session course provides an essential entry point, but achieving sustained, long-term pain management may require larger therapeutic volumes. In conclusion, a three-week course of non-invasive PRF stimulation serves as a safe, effective, non-pharmacological adjunct that aids in managing neuropathic diagnostic presentation and mitigates severe breakthrough pain experiences in patients suffering from pedal diabetic peripheral neuropathy.

## Data Availability

Data Availability: The datasets generated and analyzed during the current study are available from the corresponding author upon reasonable request and subject to data sharing agreements.

---

**Supplementary Table 1:** Unadjusted and Multivariable Logistic Regression of BPI Worst Pain Responders at 1 Month.

| Predictor | Unadjusted OR (95% CI) | p-value | Adjusted OR (95% CI) | p-value |
| --- | --- | --- | --- | --- |
| **mITT Cohort (N = 80)** |  |  |  |  |
| Treatment (Active vs. Placebo) | 3.78 (1.42-11.05) | **0.010\*** | 4.86 (1.56-17.53) | **0.010\*** |
| Age | - | - | 0.97 (0.92-1.03) | 0.369 |
| Gender (Male vs. Female) | - | - | 0.95 (0.32-2.93) | 0.933 |
| Baseline BPI Worst Pain | - | - | 1.33 (1.01-1.81) | 0.053 |
| Baseline Medication Use (Yes vs. No) | - | - | 0.38 (0.13-1.10) | 0.078 |
| **mPP Cohort (N = 60)** |  |  |  |  |
| Treatment (Active vs. Placebo) | 3.05 (1.05-9.61) | **0.046\*** | 3.65 (0.98-16.14) | 0.066 |
| Age | - | - | 0.97 (0.90-1.04) | 0.353 |
| Gender (Male vs. Female) | - | - | 0.67 (0.19-2.40) | 0.538 |
| Baseline BPI Worst Pain | - | - | 1.44 (1.04-2.06) | **0.034\*** |
| Baseline Medication Use<br>(Yes vs. No) | - | - | 0.49 (0.14-1.61) | 0.243 |

*An Odds Ratio (OR) > 1 indicates a higher likelihood of achieving responder status ( $\geq 3$ -point reduction in BPI Worst Pain) at 1 month. The unadjusted model includes only the treatment group assignment. The adjusted model controls for age, gender, baseline pain severity, and baseline medication use.*

**Supplementary Table 2A:**
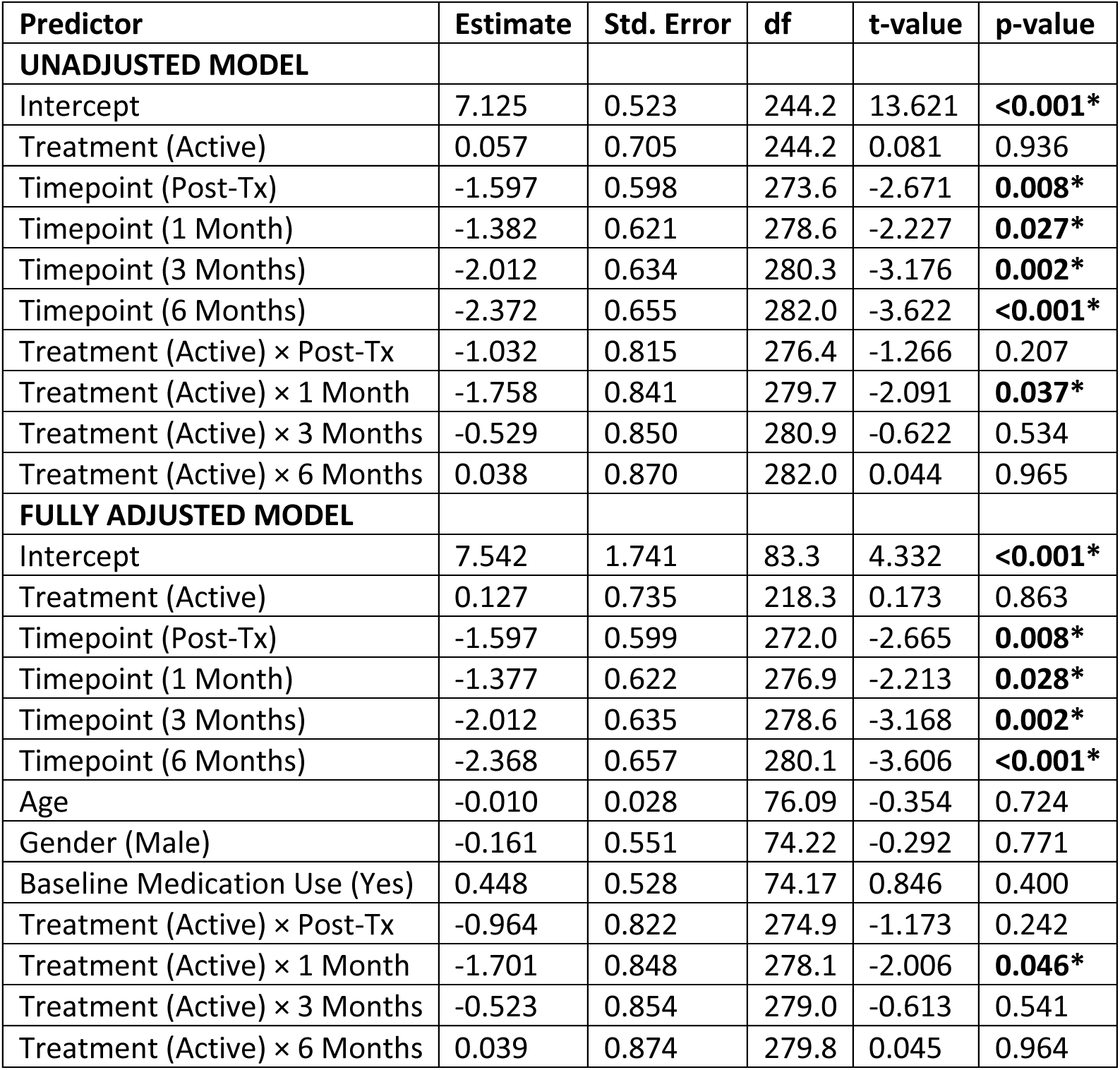
Full LMM Output for BPI Worst Pain.

**Supplementary Table 2B:** Full LMM Output for BPI Least Pain.

| Predictor | Estimate | Std. Error | df | t-value | p-value |
| --- | --- | --- | --- | --- | --- |
| **UNADJUSTED MODEL** |  |  |  |  |  |
| Intercept | 3.542 | 0.373 | 213.7 | 9.506 | **<0.001\*** |
| Treatment (Active) | -0.974 | 0.502 | 213.7 | -1.938 | 0.054 |
| Timepoint (Post-Tx) | -1.153 | 0.400 | 273.9 | -2.884 | **0.004\*** |
| Timepoint (1 Month) | -1.038 | 0.415 | 278.2 | -2.498 | **0.013\*** |
| Timepoint (3 Months) | -1.156 | 0.424 | 279.6 | -2.724 | **0.007\*** |
| Timepoint (6 Months) | -1.332 | 0.439 | 280.9 | -3.036 | **0.003\*** |
| Treatment (Active) × Post-Tx | 0.441 | 0.545 | 276.6 | 0.809 | 0.419 |
| Treatment (Active) × 1 Month | 0.324 | 0.563 | 279.4 | 0.575 | 0.566 |
| Treatment (Active) × 3 Months | 0.764 | 0.570 | 280.4 | 1.341 | 0.181 |
| Treatment (Active) × 6 Months | 0.968 | 0.583 | 281.2 | 1.662 | 0.098 |
| **FULLY ADJUSTED MODEL** |  |  |  |  |  |
| Intercept | 4.711 | 1.312 | 82.06 | 3.590 | **<0.001\*** |
| Treatment (Active) | -0.982 | 0.527 | 188.4 | -1.866 | 0.064 |
| Timepoint (Post-Tx) | -1.153 | 0.400 | 272.0 | -2.884 | **0.004\*** |
| Timepoint (1 Month) | -1.034 | 0.416 | 276.2 | -2.489 | **0.013\*** |
| Timepoint (3 Months) | -1.159 | 0.424 | 277.6 | -2.731 | **0.007\*** |
| Timepoint (6 Months) | -1.329 | 0.439 | 278.7 | -3.029 | **0.003\*** |
| Age | -0.017 | 0.021 | 76.56 | -0.798 | 0.427 |
| Gender (Male) | -0.427 | 0.418 | 74.88 | -1.022 | 0.310 |
| Baseline Medication Use (Yes) | 0.234 | 0.401 | 74.78 | 0.583 | 0.562 |
| Treatment (Active) × Post-Tx | 0.509 | 0.549 | 274.8 | 0.928 | 0.354 |
| Treatment (Active) × 1 Month | 0.395 | 0.566 | 277.5 | 0.698 | 0.486 |
| Treatment (Active) × 3 Months | 0.800 | 0.571 | 278.2 | 1.402 | 0.162 |
| Treatment (Active) × 6 Months | 0.998 | 0.584 | 278.8 | 1.710 | 0.088 |

**Supplementary Table 2C:**
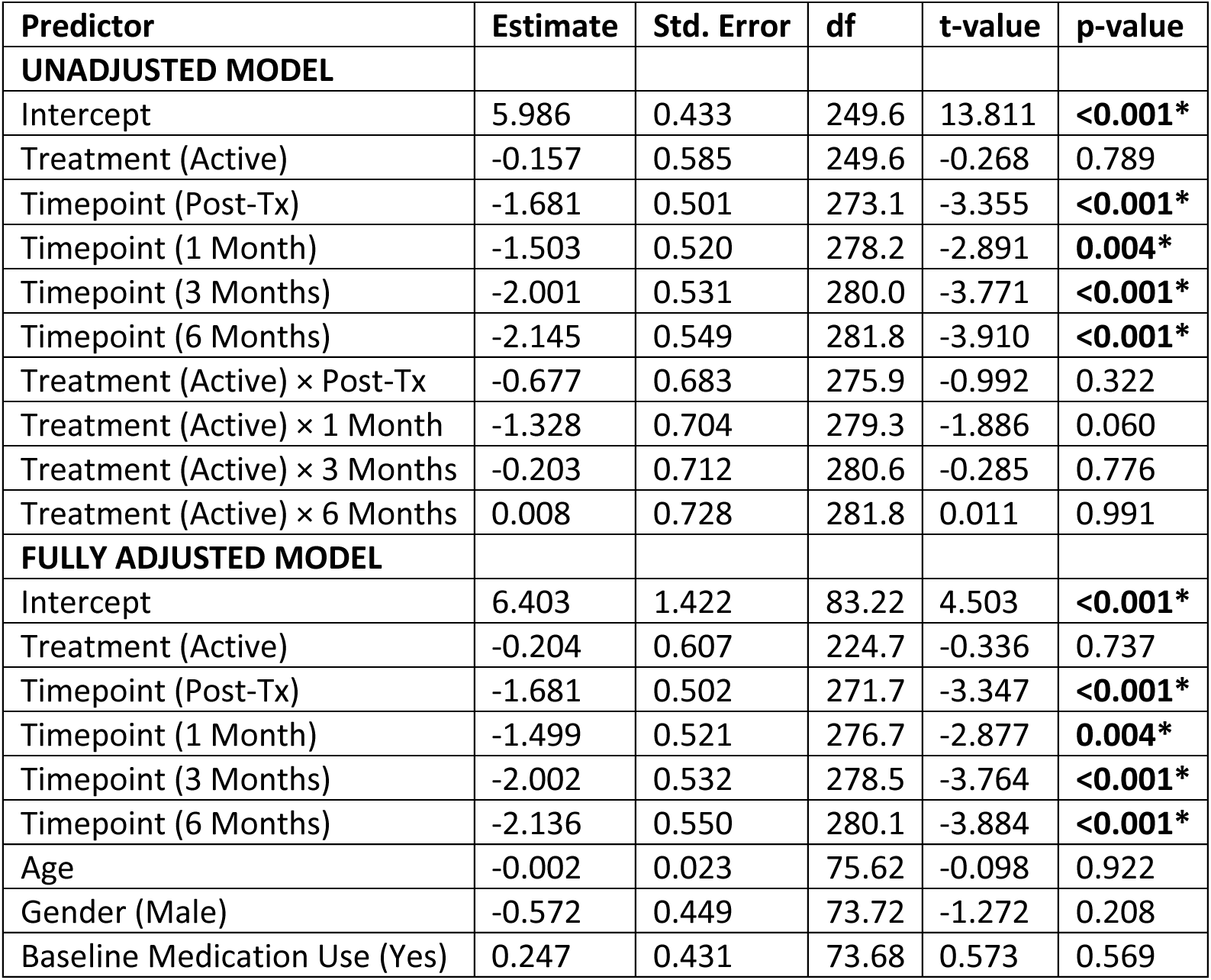
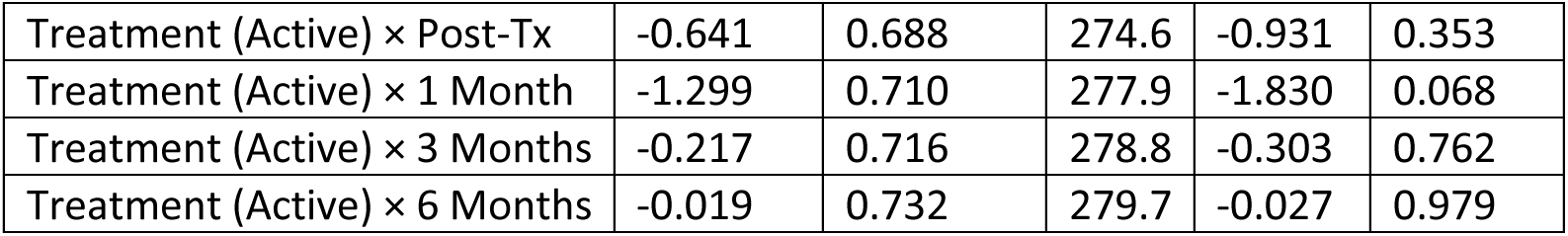
Full LMM Output for BPI Average Pain.

**Supplementary Table 2D:** Full LMM Output for BPI Present Pain.

| Predictor | Estimate | Std. Error | df | t-value | p-value |
| --- | --- | --- | --- | --- | --- |
| **UNADJUSTED MODEL** |  |  |  |  |  |
| Intercept | 3.750 | 0.477 | 233.6 | 7.859 | **<0.001\*** |
| Treatment (Active) | 0.034 | 0.643 | 233.6 | 0.053 | 0.958 |
| Timepoint (Post-Tx) | -0.431 | 0.535 | 273.2 | -0.805 | 0.422 |
| Timepoint (1 Month) | -0.513 | 0.556 | 278.0 | -0.923 | 0.357 |
| Timepoint (3 Months) | -1.134 | 0.567 | 279.7 | -2.000 | **0.047\*** |
| Timepoint (6 Months) | -0.867 | 0.586 | 281.2 | -1.478 | 0.141 |
| Treatment (Active) × Post-Tx | -1.375 | 0.729 | 276.0 | -1.885 | 0.061 |
| Treatment (Active) × 1 Month | -1.157 | 0.753 | 279.2 | -1.537 | 0.125 |
| Treatment (Active) × 3 Months | -0.395 | 0.761 | 280.4 | -0.518 | 0.605 |
| Treatment (Active) × 6 Months | -0.999 | 0.779 | 281.4 | -1.283 | 0.200 |
| **FULLY ADJUSTED MODEL** |  |  |  |  |  |
| Intercept | 4.182 | 1.615 | 81.87 | 2.589 | **0.011\*** |
| Treatment (Active) | -0.077 | 0.671 | 207.3 | -0.115 | 0.909 |
| Timepoint (Post-Tx) | -0.431 | 0.536 | 271.1 | -0.804 | 0.422 |
| Timepoint (1 Month) | -0.506 | 0.556 | 275.8 | -0.910 | 0.364 |
| Timepoint (3 Months) | -1.135 | 0.568 | 277.4 | -1.998 | **0.047\*** |
| Timepoint (6 Months) | -0.856 | 0.587 | 278.8 | -1.458 | 0.146 |
| Age | -0.006 | 0.026 | 75.29 | -0.219 | 0.827 |
| Gender (Male) | -0.611 | 0.512 | 73.49 | -1.192 | 0.237 |
| Baseline Medication Use (Yes) | 0.588 | 0.491 | 73.43 | 1.196 | 0.235 |
| Treatment (Active) × Post-Tx | -1.297 | 0.735 | 274.0 | -1.766 | 0.079 |
| Treatment (Active) × 1 Month | -1.073 | 0.758 | 277.1 | -1.416 | 0.158 |
| Treatment (Active) × 3 Months | -0.357 | 0.764 | 277.9 | -0.468 | 0.641 |
| Treatment (Active) × 6 Months | -0.977 | 0.781 | 278.6 | -1.250 | 0.212 |

**Supplementary Table 2E:** Full LMM Output for BPI Interference Composite.

| Predictor | Estimate | Std. Error | df | t-value | p-value |
| --- | --- | --- | --- | --- | --- |
| **UNADJUSTED MODEL** |  |  |  |  |  |
| Intercept | 5.103 | 0.443 | 177.7 | 11.517 | **<0.001\*** |
| Treatment (Active) | 0.084 | 0.598 | 177.7 | 0.140 | 0.889 |
| Timepoint (Post-Tx) | -1.835 | 0.438 | 270.7 | -4.195 | **<0.001\*** |
| Timepoint (1 Month) | -1.798 | 0.455 | 274.4 | -3.951 | **<0.001\*** |
| Timepoint (3 Months) | -2.182 | 0.465 | 275.5 | -4.694 | <0.001* |
| Timepoint (6 Months) | -2.175 | 0.481 | 276.5 | -4.523 | <0.001* |
| Treatment (Active) × Post-Tx | -0.508 | 0.597 | 273.4 | -0.851 | 0.396 |
| Treatment (Active) × 1 Month | -0.870 | 0.617 | 275.7 | -1.410 | 0.160 |
| Treatment (Active) × 3 Months | -0.220 | 0.624 | 276.5 | -0.352 | 0.725 |
| Treatment (Active) × 6 Months | -0.257 | 0.639 | 277.1 | -0.403 | 0.688 |
| **FULLY ADJUSTED MODEL** |  |  |  |  |  |
| Intercept | 6.745 | 1.616 | 77.75 | 4.174 | <0.001* |
| Treatment (Active) | -0.027 | 0.622 | 159.6 | -0.043 | 0.966 |
| Timepoint (Post-Tx) | -1.835 | 0.438 | 268.9 | -4.196 | <0.001* |
| Timepoint (1 Month) | -1.795 | 0.455 | 272.6 | -3.944 | <0.001* |
| Timepoint (3 Months) | -2.185 | 0.465 | 273.7 | -4.700 | <0.001* |
| Timepoint (6 Months) | -2.168 | 0.481 | 274.6 | -4.509 | <0.001* |
| Age | -0.015 | 0.026 | 73.64 | -0.556 | 0.580 |
| Gender (Male) | -1.021 | 0.517 | 72.21 | -1.974 | 0.052 |
| Baseline Medication Use (Yes) | 0.001 | 0.496 | 72.09 | 0.003 | 0.998 |
| Treatment (Active) × Post-Tx | -0.438 | 0.601 | 271.6 | -0.730 | 0.466 |
| Treatment (Active) × 1 Month | -0.803 | 0.620 | 273.9 | -1.294 | 0.197 |
| Treatment (Active) × 3 Months | -0.194 | 0.625 | 274.5 | -0.310 | 0.757 |
| Treatment (Active) × 6 Months | -0.241 | 0.640 | 274.9 | -0.377 | 0.707 |

**Supplementary Table 3:** Longitudinal Medication Use.

| **Timepoint** | **Active Cohort (%)** | **Placebo Cohort (%)** |
| --- | --- | --- |
| *Baseline* | 63.0 | 52.2 |
| *Post-Treatment* | 52.2 | 52.2 |
| *1 Month* | 37.0 | 26.1 |
| *3 Months* | 39.1 | 28.3 |
| *6 Months* | 37.0 | 23.9 |

*\*Note: Data represents the percentage of patients utilizing concurrent systemic analgesic medications at each follow-up timepoint. A statistically significant decline in medication use was observed from baseline to 6 months in both the active cohort ( $p = 0.014$ ) and the placebo cohort ( $p = 0.002$ ), as evaluated by McNemar's test for paired proportions.*

